# Policies and strategies for HPV vaccination schedule completion in immunocompromised girls, including girls living with HIV: Qualitative insights from Eswatini, Malawi, and Uganda

**DOI:** 10.1101/2025.05.06.25327127

**Authors:** Emily E Crawford, Tosin F Ajayi, Immaculate Ampeire, Michael Baganizi, Mike N Chisema, Bridget C Griffith, Lorraine Kabunga, Thuli Magagula, Lisa-Rufaro Marowa, Akachi E Mbogu, Nobuhle Mthethwa, Stella Namutebi, Bhekiwe Shongwe, Timothy Tchereni, Frehiwot Birhanu, Xolisiwe Dlamini, Fredrick Luwaga, Sonali M Patel

**Affiliations:** Global Vaccines Delivery Team, Clinton Health Access Initiative, Boston, MA, USA; Analytics and Implementation Research Team, Clinton Health Access Initiative, Boston, MA, USA; Eswatini Country Team, Clinton Health Access Initiative, Mbabane, Eswatini; Malawi Country Team, Clinton Health Access Initiative, Lilongwe, Malawi; Uganda Country Team, Clinton Health Access Initiative, Kampala, Uganda; Expanded Program on Immunization, Ministry of Health, Mbabane, Eswatini; National AIDS Program, Ministry of Health, Mbabane, Eswatini; Expanded Program on Immunization, Ministry of Health, Lilongwe, Malawi; UNEPI, Ministry of Health, Kampala, Uganda

**Author notes:** These authors are joint senior authors (Frehiwot Birhanu, Xolisiwe Dlamini, Fredrick Luwaga, Sonali M Patel).

## Abstract

Immunocompromised girls, including girls living with HIV, face significantly increased risk for HPV-linked cervical cancer and require a differentiated human papillomavirus (HPV) vaccination schedule. While World Health Organization (WHO) vaccination guidelines exist, recommending immunocompromised individuals receive at least two, if possible three doses of HPV vaccine, little is known about how country health programs implement these recommendations. This study examines HPV vaccination policies, delivery strategies, barriers, and enablers for immunocompromised girls in Eswatini, Malawi, and Uganda—countries with a high burden of HIV and cervical cancer.

A cross-sectional qualitative study was conducted through key informant interviews and focus group discussions with stakeholders from ministries of health, implementing partners, and health workers. A document review of national policies and global publications was also conducted. Data were analyzed thematically to identify common and country-specific themes.

All three countries follow the WHO’s recommendation for a two-dose HPV vaccine schedule for immunocompromised girls. However, none have fully documented or consistently implemented policies or delivery strategies to reach immunocompromised girls with additional HPV vaccination doses. Stakeholder awareness of differentiated dosing schedules and strategies to vaccinate immunocompromised girls was limited and inconsistent. Promising strategies were identified, including use of Teen Clubs and adolescent HIV clinics to deliver HPV vaccines. Barriers included stigma, limited cold chain infrastructure, unclear operational policies, and weak data systems. Enablers included trusted health provider relationships, peer mentorship, and community awareness of cervical cancer risks.

Improving HPV vaccine delivery for immunocompromised girls, including girls living with HIV, will require documenting and disseminating clear policies, integrating vaccination into HIV and adolescent health services, scaling successful strategies, and strengthening data systems to monitor coverage. Tailored, stigma-sensitive strategies are essential to ensure equitable cervical cancer prevention for immunocompromised adolescents in high HIV-burden settings.

## Introduction

### Cervical Cancer is a disease of public health importance around the world and disproportionately affects women with HIV

Human Papillomavirus (HPV) is the most common sexually transmitted infection globally[1], and it is estimated to cause 4.5% of all cancers worldwide [2,3]. In addition, HPV causes an estimated 90-100% of cervical cancers [4,5]. Cervical cancer is the eighth most common cancer globally, with 661,021 new cases in 2022, and it is the ninth leading cause of cancer deaths, with an estimated 348,189 deaths from cervical cancer in the same year [6]. The disease burden from cervical cancer is disproportionately high in low- and middle-income countries[7], and it is the largest cause of death by cancer in 37 countries (mainly LMICs)[6]. Women living with HIV (WLHIV) have an elevated prevalence of infection with the serotypes of HPV that are most likely to cause cervical cancer. In South Africa, adolescent girls living with HIV have been shown to be at higher risk of HPV infection[8]. WLHIV are also highly affected by cervical cancer [9–13], with WLHIV facing a six-fold higher risk of developing cervical cancer compared to those without HIV [14,15]. Studies have shown that WLHIV who develop cervical cancer have a worse prognosis than HIV-negative women [16].

### WHO has announced a strategy that aims to eliminate cervical cancer as a public health problem, with a focus on HPV vaccination

In 2020, WHO adopted a Global Strategy to Accelerate the Elimination of Cervical Cancer as a Public Health Problem by 2030. This strategy incorporates the "90-70-90" targets, which aim for 90% of all girls to receive HPV vaccination by the age of 15, 70% of all women to undergo high-performance screening tests at least once by age 35 and again by 45, and 90% of all pre-cancers to be treated and invasive cancer cases to be managed by 2030 [17]. It recommends that HPV vaccines be integrated into all national immunization programs, and that they reach 90% of all girls by the age of 15 by 2030, including those who are HIV-positive. In addition to this plan, WHO issued revised guidelines for screening and treatment. These guidelines emphasized that women aged 30 to 49 in the general population and WLHIV aged 25 to 49 should be given priority for HPV screening [18]. HPV vaccination is the main primary prevention intervention for cervical cancer [17]. Scaling up HPV vaccination and cervical screening worldwide could significantly reduce the incidence and mortality of cervical cancer [7,19], with WHO estimating that HPV vaccination could prevent at least a third of HPV-related cancers [20].

### HPV vaccination is safe and effective for immunosuppressed people, including people living with HIV

Clinical trials have demonstrated that HPV vaccines are safe, immunogenic, and efficacious in HIV-positive and immunocompromised individuals. Immunocompromised individuals respond well to HPV vaccines [11]. Nevertheless, immune responses to vaccination in people living with HIV are often sub-optimal. Although immune responses to vaccination improve with antiretroviral therapy (ART), they often remain lowered [21,22]. Alternative vaccination schedules, including additional doses, are required [11,21,23]. Based on this evidence, WHO recommends a two-dose vaccination schedule for immunocompromised girls, with a third dose given where possible [20].

Stigma at many levels can prevent women and girls living with HIV from equitable access to health services [24–26]; Though data is limited, women and girls living with HIV have been shown to have lower uptake of the HPV vaccine in Canada and Uganda [27,28]. These factors indicate that girls living with HIV (GLHIV) require a tailored strategy for HPV vaccination.

### Southern and Eastern Africa are disproportionately affected by HPV, cervical cancer, and HIV, signaling a need for focused HPV prevention in this region

HPV prevalence in Sub-Saharan Africa is estimated at 24%, compared to 11.7% globally. In addition, sub-Saharan Africa has the highest cervical cancer incidence rates globally [12]; East Africa and Southern Africa have the highest incidence rate of cervical cancer in the world, with region-specific incidence rates of 40.4 and 34.9 per 100,000 women, respectively, in 2022 [6]. Eswatini has the highest cervical cancer incidence rate of any country in the world, with age-standardized incidence 84.6 per 100,000 women in 2019 [29]; the country also records the highest cervical cancer mortality rate in the world [6,7,29]. Women in Malawi and Uganda have an elevated risk of cervical cancer, even when compared to other countries in the region, with age-standardized incidence 67.9 and 56.2 per 100,000, respectively in 2020 [29]. This elevated burden can be attributed, in part, to the co-occurrence of a syndemic (synergistic epidemic) between Human Immuno-deficiency Virus (HIV) and HPV [14]. For 2024, UNICEF estimates HIV prevalence of 7.5% in adolescent girls ages 10-14 in Eswatini, 2.4% in Malawi, and 2.2% in Uganda, totaling approximately 30,000 girls ages 10-14 who require a specialized schedule of the HPV vaccine in order to effectively prevent cervical cancer [30]. Prevention is particularly important in this region, given studies that show cervical cancer incidence is increasing in African countries [31].

HPV vaccine introduction in LMICs has evolved through various stages of complexity, characterized by refining strategies, policy changes, and varied experiences across different countries. These strategies have primarily revolved around defining which population will be targeted, the age cohort, delivery methods, and enhancing planning, communication, and coordination [32]. While southern African countries are making strides towards the goal of vaccinating 90% of girls against HPV, research regarding HPV vaccination for immunocompromised girls is limited.

This paper seeks to describe the policies, strategies, barriers, and enablers for vaccinating immunocompromised girls, with a focus on GLHIV, in Eswatini, Malawi, and Uganda. The hypothesis for this research study is that, despite the availability of global recommendations, there are currently limited intentional efforts to reach HIV-positive and immunocompromised girls with full HPV vaccine schedules in these countries.

## Methods

The study took a cross-sectional qualitative approach, with Key Informant Interviews (KII) and Focus Group Discussions (FGD) held with stakeholders from national and sub-national government personnel, Non-Governmental Organizations (NGO) staff, and health care providers in each of the three study countries: Eswatini, Malawi, and Uganda. A country-specific desk review was also conducted, which included peer-reviewed articles and policy documents available for each study country. Policy documents and other grey literature were obtained through web searches and requested in KII; information from these documents was used for comparison with data obtained in the stakeholder interviews and FGD.

This study was reviewed by the Clinton Health Access Initiative’s internal Scientific and Ethical Review Committee (SERC) and approved by the Eswatini Health and Human Research Review Board (EHHRRB089/2024), the Malawi National Health Sciences Research Committee (NHSRC) (24/09/4522), and the Uganda National Council for Science and Technology (UNCST) (HS4994ES).

Participant recruitment and data collection were conducted by trained CHAI personnel between 7 November 2024 and 24 January 2025. Data was collected under written informed consent using interview guides (for KII) and discussion guides (for FGDs) tailored to each country’s context. The guides were developed using the Practical, Robust Implementation and Sustainability Model (PRISM) framework to understand contextual factors at multiple levels including the external environment, organizational characteristics and individual perspectives[33]. All KIIs and FGDs were recorded with the consent of the participants. Following the interview, the research assistants completed a detailed note-taking matrix, providing in-depth answers to each question discussed, and relying on the recordings to ensure completeness and correctness of the data. All KIIs and FGDs were conducted in English.

All data was analyzed by thematic analysis using the note-taking matrices. Qualitative data analysis was assisted by ChatGPT (OpenAI, April 2024 version), a large language model, which supported the thematic analysis and synthesis of the data. Raw data was shared with ChatGPT in thematic sections, and the model was prompted to carry out thematic analysis. All outputs were reviewed, validated, and iterated on by the study team to ensure accuracy and completeness of the analyses.

## Results

In total, 69 KIIs and 18 FGDs were conducted across Eswatini, Malawi, and Uganda between November 2024 and January 2025 (Table 1). KIIs in Eswatini were conducted with stakeholders from the Ministry of Health, other government entities, and implementing partners; three FGDs were conducted, one each with nurses, rural health motivators, and community leaders. In Malawi, KIIs were conducted with stakeholders from the Ministry of Health, other government entities, and implementing partners, at national and sub-national levels (Lilongwe and Ntchisi districts) and FGDs were conducted with health care providers in three districts of Mzimba North, Mchinji and Machinga, drawn from a total of 23 health clinics, representing all the three administrative regions of the country. In Uganda, five KIIs were conducted at the national level, and 16 in Bududa, Kampala, Lira, and Rukungiri districts. Twelve FGDs were conducted with health care providers in clinics located in the same four districts.

**Table 1.** Data collection by respondent type and country.

|  | **Eswatini** | **Malawi** | **Uganda** |
| --- | --- | --- | --- |
| Data Collection Period | November 2024 | January 2025 | December 2024 |
| ***Key informant interviews*** |  |  |  |
| Government | 15 | 19 | 21 |
| Partners | 8 | 6 | 0 |
| **Total** | **23** | **25** | **21** |
| ***Focus group discussions*** |  |  |  |
| Health workers | 2 | 3 | 12 |
| Community leaders | 1 | 0 | 0 |
| **Total** | **3** | **3** | **12** |

### HPV vaccine introduction

Uganda introduced the HPV vaccine in 2015 with a two-dose schedule as recommended by WHO at that time and a third dose for immunocompromised girls (Fig 1). Malawi introduced the HPV vaccine with a two dose schedule in a phased approach starting in 2018 with national coverage achieved in 2021. Eswatini introduced the HPV vaccine in 2023 with a single-dose schedule; a second dose for immunocompromised girls was introduced in 2024. Malawi shifted from a two-dose schedule to a single-dose schedule in 2024, with a second dose for immunocompromised girls. In 2025, Uganda shifted to a single-dose schedule with a second dose for immunocompromised girls. The KII and FGD for Uganda were conducted prior to the announcement of the shift to a single dose.

**Fig 1.**
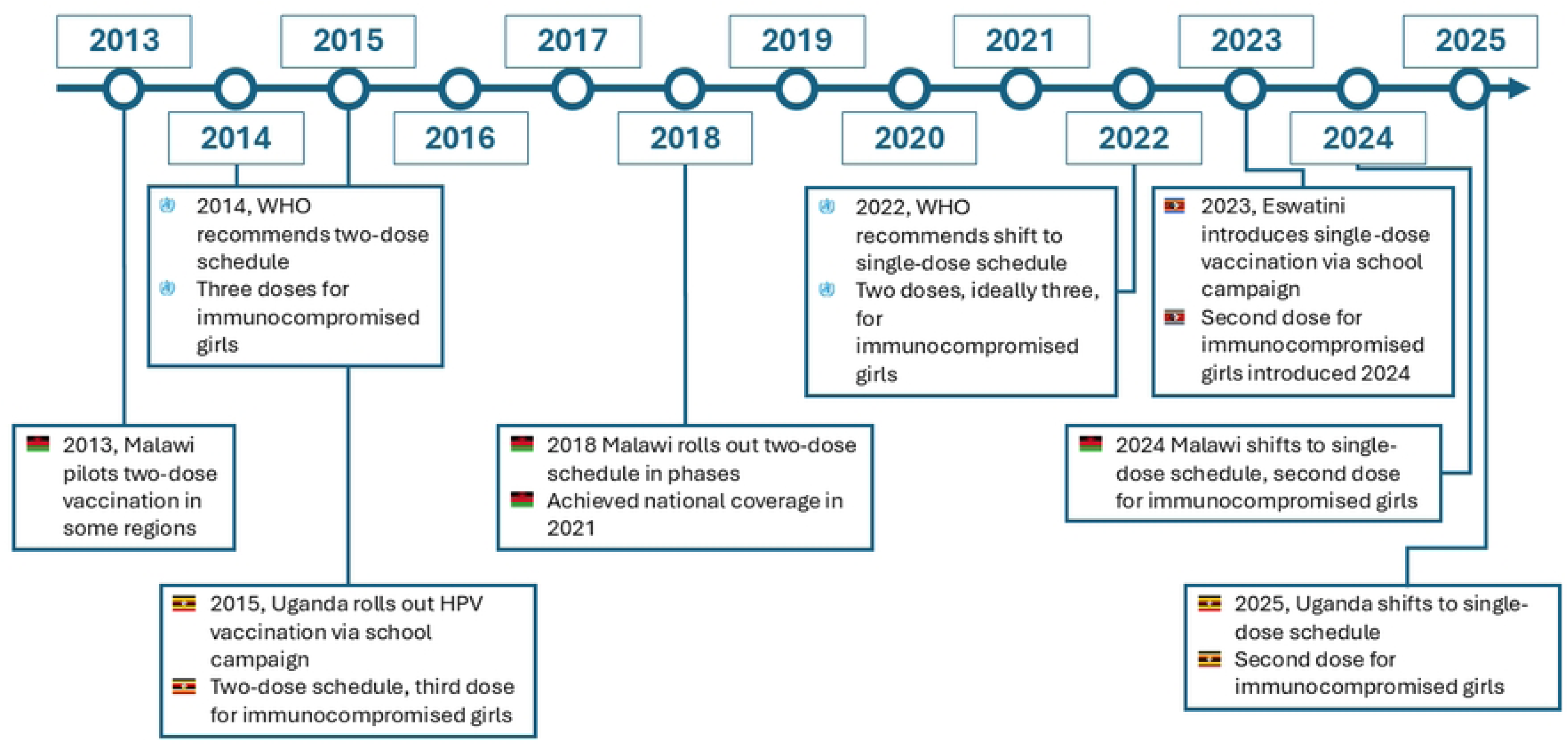
HPV vaccine introduction timeline in Eswatini, Malawi, and Uganda. Eswatini, Malawi, and Uganda all introduced the vaccine through a campaign-based delivery strategy (Table 2). Following introduction, all three countries have integrated HPV vaccination into routine health services. Malawi continues to deliver the HPV vaccine in schools as a complement to routine vaccination, and Eswatini has plans to conduct a community campaign in 2025. Uganda, in addition to routinely offering the vaccine, also delivers it in communities through Integrated Child Health Days held twice a year.

**Table 2.** HPV vaccine introduction and delivery strategies in Eswatini, Malawi, and Uganda.

|  | **Eswatini** | **Malawi** | **Uganda** |
| --- | --- | --- | --- |
| HPV vaccine introduction | 2023 | 2018 | 2015 |
| Single-dose schedule adoption | 2023 | 2024 | 2025 |
| Second dose for immunocompromised girls (year of implementation) | Yes (2024) | Yes (2024) | Yes (2025) |
| Introduction strategy | School campaign | School and community campaign | School campaign |
| Current vaccine strategy | Routine health services | Routine health services, campaigns | Routine health services, outreach |
| Target age (years) | 9-14 | 9 | 10 |
| HPV1 coverage [34] | 59% | 68% | 99% |

### HPV vaccination policy for GLHIV and immunocompromised girls

#### There is limited documentation of HPV vaccine policy for GLHIV and immunocompromised girls

While the dosing schedule for immunocompromised adolescent girls in Eswatini, Malawi, and Uganda is the same in each country - two doses at a six-month interval – documentation of this guideline varies by country. The Eswatini Vaccine Integration Framework states that the HPV vaccine should be integrated into health services, including clinics providing specific services for HIV, tuberculosis (TB) and non-communicable diseases (NCDs) [35]; although the guidelines clearly prescribe where the vaccine should be made available, the dose schedule and specific strategies for reaching immunocompromised girls are not documented in any policy or guidelines document. In Uganda, the Director General of Health Services has communicated the dosing schedule for immunocompromised girls via official letter [36], but this has not been formalized as policy. The dose schedule and strategies for reaching immunocompromised girls in Malawi are not documented in any policy or guidelines document.

#### Awareness of the dose schedule is inconsistent among stakeholders

Stakeholders, including health workers, in all three countries have inconsistent awareness of the dose schedule and strategies to fully vaccinate immunocompromised girls. Stakeholders in Eswatini know that GLHIV should receive a second dose of the vaccine, but do not have a clear understanding of how this is done in practice. Similarly, some stakeholders in Eswatini said that immunocompromised HIV-negative girls also require a second dose, but awareness of this is lower. Some respondents shared that the vaccination strategy for HIV-positive and immunocompromised girls has been intentionally kept private to avoid stigma; this strategy has led to a lack of awareness among stakeholders, affected girls, and their families about the vaccination process for GLHIV in Eswatini. In Malawi, stakeholders have low awareness of the dosing schedule for GLHIV, but they also have low awareness of the dosing schedule for HIV-negative girls. In FGDs, all health workers reported that they provide the same dose to all girls regardless of HIV status; in one health facility, respondents gave one dose to all girls, whereas in two facilities, respondents gave two doses regardless of the girl’s HIV status. In Uganda, stakeholders had low awareness of the dosing schedule for GLHIV, and many were opposed to the idea of a different schedule for this group.

### Strategies for vaccinating GLHIV

Policy, documentation, and awareness about vaccinating immunocompromised girls varied by country. Nevertheless, strategies for reaching GLHIV were similar across the three countries. In each country, some of the respondents, often health workers, were able to give examples of ways that girls with HIV are identified and given the HPV vaccine. In Eswatini, Malawi, and Uganda, the main delivery strategy for girls with HIV was delivering the HPV vaccine to girls with HIV through HIV clinics and teen clubs.

#### Strategies for vaccinating GLHIV in Malawi

In Malawi, although stakeholders were not familiar with strategies to deliver a differentiated dosing schedule to GLHIV, a small number of them were familiar with strategies for identifying and vaccinating GLHIV. Some clinics reportedly offer HPV vaccination to girls attending ART clinics, although it was said that HPV records do not state HIV status. In some communities, community health workers conduct door-to-door outreach to identify eligible girls for vaccination; in some cases, they may also identify GLHIV.

The strategy that was described in the most detail in one FGD in Malawi was delivering the vaccine at Teen Clubs, which are support groups for adolescents with HIV and the place where many adolescents receive their ART. Respondents in one FGD clearly described their process of conducting health talks about the HPV vaccine with information targeted specifically to their members, framing the vaccine as a routine preventive measure, and administering the vaccine during the same session. Other adolescent-friendly health services, such as family planning, are also offered at the Teen Club meetings. Peer mentors also operate in the club, serving as positive role models for vaccine acceptance for other members of the group. The health workers conduct concurrent health discussions with parents during the Teen Club meetings, which promotes vaccine uptake.

#### Strategies for vaccinating GLHIV in Uganda

Uganda was similar to Malawi in that a differentiated vaccination schedule was not offered to GLHIV, but some strategies are used to administer the HPV vaccine to GLHIV. Some ART clinics offer youth-friendly services, and some have linkage facilitators that ensure GLHIV are connected to health care. One health worker also gave an example of an ART clinic scheduling appointments for vaccination when adolescent girls come for their refills. It is not clear, however, whether any ART clinics currently offer HPV vaccination.

#### Strategies for vaccinating GLHIV in Eswatini

The strategies to reach GLHIV with two doses of the HPV vaccine are clearer in Eswatini. Although awareness of strategies for vaccinating immunocompromised girls was not consistent across all stakeholders, some Ministry of Health (MOH) personnel and health care providers were able to describe existing practices for identifying and administering two doses of HPV vaccine to GLHIV. These respondents described training and supportive supervision for health workers in ART clinics to administer the HPV vaccine in the HIV clinic. Some respondents mentioned integration of HPV vaccination in other areas of the hospital for girls who have other immunocompromised conditions, such as cancer or diabetes. However, most responses focused on GLHIV.

Within the ART clinics, identification of girls for vaccination varies. Some GLHIV attend the clinic regularly to receive their ART; during these visits, their nurse suggests they receive the HPV vaccine. In other cases, clinics coordinate with Teen Clubs, where they identify and/or vaccinate youth who are in attendance. Some nurses in Eswatini described using the Client Management Information System (CMIS) to identify girls who are living with HIV for HPV vaccination. In some communities, community health workers (CHWs) are able to speak to GLHIV in their homes to educate them about the need for a second dose. For all GLHIV, the second dose is recorded in their HIV patient card rather than their HPV vaccination booklet to avoid stigma.

#### Strategies for vaccinating immunocompromised girls are similar across study countries

Across the three study countries, the main strategies for delivering HPV vaccine to immunocompromised girls was through ART clinics and Teen Clubs. Strategies for vaccination focused on reaching GLHIV in all three countries, and in both Malawi and Uganda, no strategies were mentioned for reaching HIV-negative immunocompromised girls.

Carrying out vaccination at the HIV or ART clinic has advantages and disadvantages. Respondents in Eswatini say that this approach is helpful because it allows girls to receive the vaccine from a healthcare provider whom they have a relationship with, which helps with trust and uptake. However, most HIV clinics do not have cold storage available for vaccines, so clinics must make special arrangements and adaptations to ensure appropriate vaccine storage. Respondents with knowledge of different health facilities described different approaches, including storing vaccines in the EPI department of facilities that have them, temporarily storing vaccines in the maternity unit on HIV clinic days, and storing vaccines in cooler boxes during HIV clinic hours.

Carrying out vaccination through Teen Clubs has many of the same advantages and disadvantages as delivery in HIV or ART clinics. Teen Clubs are a place that encourage vaccine uptake as there is peer support, and there is little risk of stigma as all the participants are HIV-positive. However, disadvantages of delivery at Teen Clubs include the same lack of cold chain storage that clinics experience, as well as sporadic meeting dates and transportation challenges for GLHIV who have low access to transportation.

### Barriers for HPV vaccination among GLHIV

#### Policy and strategy for fully vaccinating GLHIV are not clear

As described above, a key barrier to fully vaccinating GLHIV is a lack of clear policy and strategy for delivering the HPV vaccine to this population. This lack of policy and strategy leads to low knowledge amongst health care providers and low levels of service delivery to this target group.

#### HIV-related stigma is identified as an important barrier to HPV vaccination

HIV-related stigma was named as an important barrier to HPV vaccination in all three countries. Difficulty speaking about HIV status leads to lower health worker knowledge about differentiated dosing schedules and difficulty identifying GLHIV. It also leads to lower levels of awareness amongst parents of GLHIV that their daughters require a second dose of the vaccine.

#### Stigma leads to challenges in disclosing HIV status, complicating access to differentiated care

Stigma leads to challenges related to HIV disclosure as well. In Eswatini, not all GLHIV are told their HIV status by 9 years, the age of vaccine initiation. Administering two doses of the vaccine to GLHIV requires careful explanation to avoid inadvertent disclosure. This barrier is even more defined in Uganda, where the age of HIV disclosure is 13, whereas the age of HPV vaccination is 10. In addition, some health systems do not disclose patient HIV status to all health workers, particularly to Community Health Workers.

#### GLHIV require tailored health communications around HPV vaccination

A lack of tailored messaging around HPV vaccination for GLHIV is another important barrier. Stakeholders in Eswatini reported that GLHIV and their parents often do not trust health messaging unless it comes from a girl’s nurse at the ART clinic. They also said that GLHIV struggle with treatment fatigue and may resist vaccination due to a feeling that they are constantly subject to new treatments and initiatives. This lack of specific health messaging for GLHIV also leads to low awareness for GLHIV and their parents of their need for a second dose of the vaccine, limiting demand.

#### HIV-specific data makes tracking HPV vaccination for GLHIV difficult

Health system weaknesses were another commonly reported barrier. In Malawi and Uganda, stakeholders spoke to a lack of data – specifically that HMIS and paper tools for HPV vaccination do not indicate HIV status. Health workers also spoke of stockouts for paper tracking tools for HPV vaccination in health facilities and for Community Health Workers. These gaps can prevent identification of GLHIV who require a second vaccine dose, can stop GLHIV and their parents from receiving relevant information about their health needs, and lead to poor tracking of vaccine coverage amongst GLHIV. Respondents in the same two countries reported limited provider knowledge, limited availability of Teen Clubs, and transportation challenges for girls to attend teen clubs. An additional barrier is a lack of cold storage for HPV vaccines in points of care for GLHIV, including ART clinics, Teen Clubs, and outreach activities.

#### GLHIV also experience vaccine barriers unrelated to HIV

Respondents also identified barriers to HPV vaccination that affect all girls, not only GLHIV. Inconsistent vaccine supply chain, which leads to stockouts, was the most commonly named barrier to vaccination in all three countries. Lack of clear policy for HPV vaccination, and lack of coordination across health system departments, and weak financing were also key barriers. In Malawi, the overall lack of provider knowledge about the appropriate dosing schedule was another important barrier.

Community misconceptions and misinformation as a barrier to HPV vaccination was also commonly reported across the three countries. Respondents say that many people believe the vaccine is a method of contraception, a cause of infertility, or even a way to kill adolescent girls. The belief that the vaccine is satanic or a form of devil worship was also reported in all three countries. In Malawi and Uganda, respondents also reported that parents of GLHIV fear that the vaccine could harm their child or reduce the effectiveness of their ART. In the same countries, some people, including health workers, confuse HIV and HPV, leading to confusion around what the HPV vaccine is for.

### Enablers for HPV vaccination among GLHIV

#### HIV-specific service availability provides a platform for vaccine delivery

Respondents also spoke to enabling factors for vaccinating GLHIV. HIV-specific services for adolescent girls, including established ART clinics and teen clubs, are an existing platform in Eswatini, Malawi, and Uganda that can support differentiated HPV vaccine delivery in these groups. Respondents indicated that this specific care is already common in Eswatini and has potential for scale-up in Malawi and Uganda. The health systems in Eswatini and Uganda have guidelines to integrate HPV vaccination, and in Eswatini this guidance specifically speaks to HIV clinics. The Eswatini health system has additional support, as health workers have received training on delivering the HPV vaccine to GLHIV and data systems can monitor HPV vaccination by HIV status.

Respondents in Eswatini and Uganda cited positive relationships between GLHIV and their health care providers as a promoter of vaccine uptake. In Malawi, community outreach and peer support were also named as enablers. In Uganda, family support groups, peer support strategies, and adolescent clinics were also named as existing structures that could support HPV vaccination for GLHIV.

#### Cervical cancer awareness promotes vaccine uptake

A commonly named enabler for HPV vaccination in all three countries was community awareness of cervical cancer. In Eswatini and Malawi, stakeholders credited effective communications campaigns with creating demand for cervical cancer prevention through HPV vaccination. In all three countries, respondents said that a fear of cervical cancer and a desire to prevent it acts as a key driver in vaccine uptake. Furthermore, health system stakeholders in the study countries have strong knowledge about HPV vaccination and are largely committed to its implementation.

## Discussion

This study is an initial effort to describe how the HPV vaccine is delivered to GLHIV, particularly in countries with a high burden of both HIV and cervical cancer. We used KIIs and FGDs to generate novel insights into the policies for fully vaccinating immunocompromised girls for HPV in Eswatini, Malawi, and Uganda, the extent to which each country implements strategies to deliver the vaccine to these girls, and the associated barriers and enablers A differentiated HPV vaccine schedule for immunocompromised girls, including GLHIV, is essential; multiple studies have investigated the most appropriate dosing schedules as well as vaccine effectiveness for these groups [20–22]. However, there is almost no literature on how health systems can effectively deliver the recommended vaccination schedule to immunocompromised girls.

### Cross-Country Policy Gaps and Missed Opportunities

While all three countries have adopted WHO’s recommendation for a two-dose HPV schedule for immunocompromised girls, including GLHIV, policy clarity and operational guidance vary substantially. In Eswatini, the Vaccine Integration Framework references incorporating HPV vaccination into services for HIV, TB, and non-communicable diseases, but it does not include specific guidance on dosing schedules or implementation strategies tailored to immunocompromised girls [35]. In Uganda, a letter from the Director General of Health Services communicated the dosing schedule for GLHIV, but this guidance has not been formally adopted as national policy [36]. In contrast, Malawi’s policy documents lack any reference to differentiated HPV vaccination for immunocompromised girls. These gaps, which were identified through KIIs but confirmed with documents obtained in the desk review, have led to uncertainty among frontline health workers regarding who should receive a second dose and how delivery should be managed [32,39].

This absence of formal documentation not only undermines consistent practice but also limits opportunities for provider training, supervision, and the allocation of necessary resources. In systems where HPV vaccination is delivered through multiple platforms— schools, health clinics, and outreach programs—lack of a clear, unified policy exacerbates confusion and opens the door to inequities in access. The challenge is further compounded for GLHIV, who may experience stigma or late HIV disclosure that interferes with their ability to access a tailored vaccination schedule. As shown in this study and echoed in prior research, implementation success relies not only on adopting WHO guidance [20] but on translating it into clear, operational policy and ensuring that it is communicated throughout all levels of the health system [10,27](Chambers et al., 2022; Okoye et al., 2021).

#### Barriers to complete vaccination of GLHIV

The health systems and health facilities in Eswatini, Malawi, and Uganda share similar barriers to HPV vaccination for GLHIV. Many of health systems barriers identified in this study are also documented in the literature, including stockouts, weak health worker knowledge and training, and insufficient data [32,39–41]. Another critical health systems barrier identified in this study is a lack of cold storage for vaccines; though the scope of this study does not allow estimation of how widespread this challenge is, it was frequently mentioned in KIIs. While these issues apply to HPV vaccination programs broadly, they are magnified when applied to GLHIV. Awareness and implementation of vaccination for GLHIV was uniformly lower than awareness of general HPV vaccination strategies in the respondents to this study. Limited availability of scientific literature regarding implementing HPV vaccination for GLHIV highlights this equity gap for health service delivery.

Findings from this study reinforce existing literature on community-level barriers to HPV vaccination. Misinformation and misconceptions—such as beliefs that the vaccine causes infertility, has harmful side effects, or is ineffective—were frequently reported by participants and are well-documented in prior studies [40–42]. Respondents also highlighted the influence of gender dynamics, echoing research by Deignan et al. [42], which found that gendered health-seeking behaviors, male dominance in household decision-making, and skepticism about vaccines targeted exclusively at girls often hinder uptake in sub-Saharan Africa. These community-level challenges are amplified for GLHIV, who face added barriers due to HIV-related stigma and lower access to tailored health information. As Huff et al. [43] note, individuals with HIV are often unaware that HPV vaccination is recommended or beneficial for them—an insight that was echoed by several respondents in this study. Together, these findings underscore the need for stigma-sensitive, gender-informed community engagement strategies to address both general misconceptions and barriers specific to GLHIV.

### Enablers for HPV vaccination for GLHIV

Eswatini, Malawi, and Uganda are all taking steps towards implementing a vaccine program aligned with WHO recommendations that will reach GLHIV. Existing infrastructure for delivering HIV-specific services to adolescent girls and the existence of teen clubs and adolescent-friendly ART clinics supports delivery of the HPV vaccine and provides the building blocks for scale-up of HPV vaccine delivery to GLHIV in the areas of Malawi and Uganda that have not already done so. This study showed that the long-term relationship between GLHIV and their ART provider is an enabler for vaccine uptake. Peer support to promote HPV vaccination was also seen as an enabler in this study and another study in GLHIV in Uganda [28]. Another important enabler for HIV vaccination across the three countries is awareness of the risks of cervical cancer, which has been observed in other qualitative work in these countries[44].

#### Promising Strategies for HPV Vaccination for GLHIV

Despite policy and operational gaps, this study identified strategies in all three countries that are currently being used to deliver the HPV vaccine to GLHIV and have potential for scale-up. Among these strategies are Teen Clubs and adolescent-friendly ART clinics, which serve as key touchpoints for ongoing HIV care [28,37]. These settings offer trusted relationships between health workers and adolescent girls and have been successfully leveraged to deliver HPV vaccination in some contexts. In Malawi, for example, providers at health facilities reported using tailored health talks at Teen Clubs to introduce the vaccine, administering it during ART refill visits, and engaging peer mentors to encourage uptake. The integration of other adolescent health services, such as family planning, further normalizes vaccination and increases service acceptability. These findings align with other literature showing the effectiveness of peer-led, adolescent-centered HIV care in promoting vaccine uptake [28].

In Eswatini, efforts to deliver HPV vaccination through HIV clinics appear more developed. Respondents described structured training for ART clinic staff, supportive supervision, and data tools like the Client Management Information System (CMIS), which help identify GLHIV due for vaccination. Community health workers (CHWs) were also involved in outreach and follow-up for second doses. Although logistical issues like cold chain storage were raised, health facilities have adapted by using cooler boxes or coordinating with maternity wards or EPI units to store vaccines on ART clinic days. These integrated delivery models demonstrate the feasibility of combining services and may offer replicable models for other countries with high burdens of HIV and cervical cancer. This type of service integration is likely to be feasible across other high-burden HIV and cervical cancer countries and is currently being tested in Zambia[38].

### Study Limitations

This study used qualitative methods to understand policies, strategies, barriers, and enablers for HPV vaccination for immunocompromised girls, including GLHIV While this allowed for rich, contextual insights, it limits generalizability. Stakeholder responses were occasionally conflicting, and interpretation relied on triangulation by country-based research team members. Additionally, although this study aimed to examine strategies for all immunocompromised girls, most stakeholder responses referred only to girls with HIV, the largest group of immunocompromised girls in the study countries.

#### Implications for Policy and Practice

The findings of this study point to several critical actions needed to strengthen HPV vaccine delivery for immunocompromised girls, particularly GLHIV, in Eswatini, Malawi, and Uganda. These findings are likely applicable to other health systems, particularly those in LMICs with a high burden of HIV among adolescent girls. While national adoption of WHO’s recommended two-dose schedule for immunocompromised girls represents important progress, effective implementation will require intentional, equity-focused design and operational planning. This study also shares insights into barriers and enablers for life-course vaccination and differential service delivery.

#### Develop and Disseminate HPV Vaccination Policies for GLHIV

A priority for all three countries is the development and dissemination of clear, national policy documents that explicitly define the HPV dosing schedule and delivery strategies for immunocompromised girls, including GLHIV. These guidelines should be developed collaboratively with personnel from departments that focus on immunization, HIV, adolescents, and girls. They should describe delivery strategies across service points— particularly HIV clinics, Teen Clubs, and adolescent-friendly services—and be supported by accompanying training, supervision tools, and job aids for frontline providers. Evidence from similar policy implementation efforts highlights the importance of clear documentation and multilevel dissemination in improving service consistency and health worker confidence [10,27,32,45].

#### Strengthen and Scale Up Existing HIV Service Delivery Mechanisms

HIV care infrastructure, especially Teen Clubs and ART clinics, provides a high-potential platform for integrated HPV vaccine delivery. Countries should scale up successful practices already in use—such as peer mentorship, synchronized vaccine and ART delivery, and use of adolescent-friendly messaging—and embed them in standard operating procedures across HIV and immunization programs. Where promising models exist, such as the integration of HPV services in ART clinics in Eswatini or Teen Clubs in both Eswatini and Malawi, countries should invest in structured learning exchanges to share best practices, identify gaps, and develop contextually appropriate adaptations[28,35].

Cold chain limitations in HIV clinics must also be addressed through innovative strategies, such as scheduled delivery days with cooler boxes or interdepartmental coordination, to ensure consistent vaccine availability. Health workers delivering HIV care should be trained to offer HPV vaccination alongside routine services, reducing missed opportunities for this high-risk group.

#### Strengthen HPV Data and Monitoring Systems for GLHIV

This study highlights the gap in measurement of HPV vaccine coverage by HIV status. Adapting data systems to capture, disaggregate, and report on coverage for immunocompromised groups—particularly GLHIV—is essential for identifying gaps and holding systems accountable [4,24,39]. Tools like the Client Management Information System (CMIS) in Eswatini can serve as examples for adaptation. Integration of HPV vaccination into ART registers, patient-held cards, and digital health tools should also be explored. SMS reminder systems, shown to be feasible in Malawi and already a capability of the Eswatini CMIS, could improve second-dose completion if integrated into immunization data systems and funded[41,46].

#### Reach HIV-Negative Immunocompromised and Other Marginalized Girls

While GLHIV are a well-defined target population within the health system, HIV-negative immunocompromised girls—such as those with cancer, diabetes, or autoimmune conditions—may be harder to identify and vaccinate. In alignment with the principles of universal health care, strategies should be developed to ensure equitable access to care for these girls [47]. Approaches may identify these girls through specialized care units or through linkage with NCD programs as advised in Eswatini’s Vaccine Integration Framework[35]. Additional outreach strategies are needed for other marginalized populations, including girls who live far from clinics, are newly diagnosed with HIV, migrate for work, or are not currently in HIV care. These strategies should be developed in collaboration with organizations already working with these groups and based on principles of equity and community engagement [20].

#### Promote Stigma-Sensitive, Gender-Aware Community Engagement

Stigma around HIV and gender norms were identified as central barriers to HPV vaccination for GLHIV. National programs should co-design communication strategies with adolescents, caregivers, peer mentors, and healthcare providers to ensure messaging is accurate, inclusive, and sensitive to privacy concerns. This includes carefully framing HPV vaccination for GLHIV as part of routine preventive care rather than a separate, stigmatized intervention. Building on strong community awareness of cervical cancer prevention—an enabler identified across countries—can help shift the narrative toward protection and empowerment [42–44].

## Conclusion

This study reveals that while HPV vaccine strategies for immunocompromised girls exist in principle, intentional design and implementation are largely lacking. Countries can leverage existing HIV service platforms, adolescent-friendly infrastructure, and cervical cancer awareness to close this equity gap. Expanded Program on Immunization (EPI) and HIV programs should collectively work together to design targeted vaccination strategies for GLHIV. Formalizing, resourcing, and monitoring strategies for vaccinating immunocompromised girls is essential for cervical cancer prevention in high HIV-burden settings.

## Data Availability

The qualitative data for this study were collected using structured note-taking matrices rather than full transcripts. These de-identified matrices contain potentially sensitive information and are not publicly available. However, de-identified data for each country included in the study may be made available upon reasonable request to the corresponding author and subject to approval from the relevant national ethics committee in each country.

## Acknowledgments

We thank the Ministries of Health in Eswatini, Malawi, and Uganda for their support and collaboration. We also acknowledge the contributions of field researchers and data collection teams in all three countries, as well as the study participants—including health workers, policy-makers, and community leaders—whose insights informed this work.

In Malawi, we thank Tuweni Chumachapera and Dr. Henry Phiri from the Ministry of Health, and Andrews Gunda from the Clinton Health Access Initiative (CHAI). In Eswatini, we thank Nyasatu Ntshalintshali and Nomfundo Mncina from CHAI. We also acknowledge Jessica Gu, Shadrack Mngemane, and Laure Anais Zultak from CHAI’s Global Vaccines Delivery team.

ChatGPT (OpenAI, April 2024 version) was used to assist with qualitative data analysis and editorial support for the first draft of the manuscript. All AI-assisted content was reviewed and refined by the study team.

